# Addressing the Needs and Identifying Supports for Parents of Chronically Ill Adolescents and Young Adults in their Shared Transition from Pediatric to Adult Care: A Scoping Review Protocol

**DOI:** 10.1101/2025.01.17.25320750

**Authors:** Pranshu Maini, Claudia Tersigni, Samantha Micsinszki, Sarah Cairns, Kate Murray, Karen Frost

**Affiliations:** Faculty of Health Sciences, McMaster University, Hamilton, ON, Canada; Schulich School of Medicine and Dentistry, Western University, London, ON, Canada; CanChild Centre for Childhood Disability Research and the School of Rehabilitation Science, McMaster University, Hamilton, ON, Canada; Health Sciences Library, McMaster University, Hamilton, ON, Canada; Robbie’s Rainbow, Oakville, ON, Canada; Division of Gastroenterology, Hepatology and Nutrition, The Hospital for Sick Children, Toronto, ON, Canada

**Keywords:** adolescent and young adults, AYAs, chronic illness, parent, parental support, parental preparedness, transition to adult care, transition resources, youth, scoping review

## Abstract

**Introduction:** The transition from pediatric to adult healthcare marks a pivotal period for chronically ill adolescents, as they transition from a highly supportive and family-oriented environment to an adult-, and a more individual-oriented healthcare system that places a greater emphasis on personal responsibility and independence. Parents, given their firsthand experience managing their child’s healthcare, play a central role in ensuring a smooth and successful transition, yet their perspectives on the barriers and facilitators of this complex process remain vastly underexplored. This scoping review aims to assess and provide comprehensive insights into parents’ perceptions of the successes and challenges during their adolescents’ transition from pediatric to adult healthcare.

**Methods & analysis:** This scoping review is led by patient partners and will be guided by the Peters et al. and the Joanna Briggs Institute (JBI) guidelines for scoping reviews. The preliminary search strategy will be developed and calibrated in Ovid MEDLINE and will be subsequently replicated in CINAHL, PsychInfo, Embase, Web of Science, and Sociological Abstracts from inception through December 18th, 2024, including all types of studies. Grey literature sources recommended by patient partners and clinical and qualitative research experts will also be included. Two reviewers will independently perform the title and abstract review of all studies against the pre-defined inclusion and exclusion criteria, followed by the full-text review of included studies. The reference list of all included studies will also be screened to maximize the retrieval of relevant sources. Data will be extracted and analyzed quantitatively and qualitatively, with the study procedural and reporting format following PRISMA-ScR guidelines.

**Ethics & dissemination:** This scoping review, through the broad and systematic mapping of existing literature, aims to provide a foundation for developing targeted support systems, strategies and interventions to address the unique needs and barriers faced by parents and caregivers of chronically-ill adolescents during this critical transition to adult care.

**Strengths and limitations of this study:**

- The inclusion of scholarly, peer-reviewed and grey literature across 6 specialized and cross-disciplinary databases will help ensure a comprehensive coverage of relevant literature, minimizing publication bias, enhancing data reproducibility and methodological rigor.
- The involvement of patient partners in driving this review from conceptualization and planning to data synthesis and reporting of results will help ensure that the research question and interpretations of findings are grounded in patient priorities, ultimately enhancing the review’s relevance and applicability.
- Findings from this scoping review will be extended to inform subsequent phases of a national multicentre study that aims to enhance parental preparedness of parents and caregivers of chronically-ill adolescents to better support their transition from pediatric to adult care.
- In employing a practical and focused strategy, the inclusion of grey literature solely recommended by patient partners (i.e. patients and caregivers/family members) and clinical and research experts (i.e. pediatric and adult gastroenterologists, nurse practitioners, and clinical and qualitative researchers) may introduce selection bias by excluding lesser-known yet potentially impactful sources.
- The inclusion of literature only published in English may limit generalizability of findings to non-English-speaking populations and unique cultural contexts and healthcare settings.

## Introduction

Healthcare transition (HCT) is defined as ‘the purposeful and planned movement of adolescents and young adults with chronic, long-term physical and medical conditions from pediatric to adult-oriented healthcare systems.’^1^ As individuals requiring specialized care, lifestyle adaptations, and extensive coordination across multiple facets of healthcare, HCTs can pose a major challenge for adolescents and young adults (AYAs) especially amidst life’s competing priorities and the growing demands of emerging adulthood.^2,3^ This process also represents a complex and pivotal juncture for parents and caregivers of these chronically ill adolescents.^2,3,4,5^

Parents, in hopes of instilling a sense of stability and security, often instinctively offer adolescents their unwavering support in alleviating some of the cognitive and emotional strain associated with managing their health in tandem with their expanding life commitments.^6^ Yet, adolescents often remain heavily reliant on parental support due to their limited social experiences and autonomy, a characteristic which may persist into early adulthood and inadvertently hinder the development of key self-management skills and age-aligned competencies.^7,8,9^ For example, adolescents with inflammatory bowel disease (IBD), an umbrella term used to describe chronic inflammatory conditions of the gastrointestinal tract, namely Crohn’s disease and ulcerative colitis, compared to their healthy peers, typically achieve fewer psychosocial developmental milestones, such as participating in unsupervised trips, holding part-time jobs during school, and infrequently attend large social gatherings, underscoring their delayed progression towards adulthood.^10,11^ On the contrary, parents develop an intimate understanding of their child’s medical history, treatment regimens, and individualized needs, which positions them as an instrumental support system during this transition, although leaving them challenged to balance their roles as protectors and promoters of independence.^12,13,14^ Therefore, when parents or caregivers lack the appropriate support, resources, or guidance in managing their own transition to a less central caregiving role, the foundations of the family’s shared preparedness may falter.

“Parental preparedness” encapsulates the capacity of parents to foster self-management and autonomy in adolescents as they approach adult care.^15,16^ However, while adolescents’ transition to adult care is often supported by a wide array of resources and systems, there remains a paucity of literature and resources aimed at enhancing parents’ readiness to transition. Studies have previously reported that parental stress and anxiety during this transition can affect adolescents’ disease management, contributing to increased healthcare utilization and potentially adverse health outcomes.^17,18,19,20,21,22,23^ This empirical evidence emphasizes the importance of offering tailored and targeted support for parents to facilitate a healthier and more effective transition for these caregivers and their adolescents.

This scoping review forms Phase 1 of a national, 3-phase multi-centre study titled “Parental Resource Offering Active Care Transition for Adolescents with IBD,” also known as the “PROACT-IBD” Study that aims to enhance parental preparedness for parents of youth with IBD and raise awareness of their pivotal role using an exploratory sequential mixed methods approach rooted in narrative medicine. More specifically, this scoping review is designed to systematically identify the unique needs, experiences, and barriers faced by parents and caregivers of chronically-ill adolescents to generate actionable recommendations, tailored support systems, and resources to strengthen parental preparedness during this HCT. Ultimately, this foundational work will help inform subsequent phases of the study and ensure that research processes are grounded in a thorough understanding of parental needs and perspectives.

## Methods

### Design

This protocol describes a scoping review guided by Arksey and O’Malley’s five-step framework, the revised recommendations made by Peters *et al*., and the Joanna Briggs Institute (JBI) Manual for evidence synthesis.^24,25^ As per the sixth stage in the JBI manual for scoping reviews, the core elements of consultation with relevant stakeholders and/or experts, such as healthcare providers, researchers, and parent/patient partners are being prioritized and integrated into the review from the outset.^25^ The study design and research strategy will be developed and further strengthened with the support of an expert librarian consultant at McMaster University. The flow and reporting of data will follow the Preferred Reporting Items for Systematic reviews and Meta-Analyses extension for Scoping Reviews (PRISMA-ScR) guidelines.^26^ To ensure transparency and consistency in documenting patient and public involvement, the reporting of patient engagement activities will be in accordance with the Guidance for Reporting Involvement of Patients and the Public 2 (GRIPP2) short form.^27^ This protocol has been reported using the PRISMA-ScR checklist (online supplemental appendix 1) and the GRIPP2 short form (online supplemental appendix 2).^26,27^

### Review team

This review is being led by patient partners and advocates in close collaboration with a multidisciplinary team comprised of expert clinicians and academics in the field of pediatric gastroenterology and inflammatory bowel disease (PM, CT, SM, KM, KF), a librarian consultant (SC), and a postdoctoral research fellow (SM). The team also reflects multi-professional expertise in nursing (SM, KF), knowledge translation and mobilization (PM, CT, SM, KM, KF), and lived experience expertise in navigating the transition from pediatric to adult healthcare (PM, CT, KM).

### Patient and public involvement

Uniquely, this review is driven primarily by patient partners and advocates having previously navigated the healthcare transition (HCT) either as patients or as parents/caregivers of adolescents with IBD. With extensive research experience, all patient partners (PM, CT, SM, KM), actively led each stage of the research process, from conceptualization and planning to protocol development and write-up, and will continue to be involved in subsequent phases, including study selection and screening, data charting and extraction, data analysis, and dissemination of results (i.e. manuscript write-up). At the study’s inception, the team also collectively developed and briefed on a charter of roles and responsibilities to ensure clarity of goals, expectations, and preferred methods of communication.

### Stage 1: Identifying the research questions

The primary aim of this review is to explore parents’ perceptions of the HCT experience, describing successes and challenges influencing parental preparedness in navigating this shared transition with chronically ill adolescents and young adults (AYAs). Findings from this study will provide valuable insights into the role parents and caregivers undertake in the transition from pediatric to adult care, while informing the growing need for parent-focused support and resources to ensure a smooth and well-coordinated transition for parents, patients, and families.

The primary research question this review aims to address is:

What are parents’ perceptions of the successes and challenges and support needs in navigating the shared transition from pediatric to adult care with their chronically ill adolescents?

Additionally, this review will address the following secondary questions:

1. What is the current landscape of support and preparedness for parents and caregivers of chronically ill AYAs during HCT?
2. What types of studies exploring parental preparedness in HCTs have been previously conducted (e.g., quantitative, qualitative, or mixed-method methodology)?
3. Which specific types of parental support and resources have been explored in prior research involving HCTs?
4. What parent-focused tools, supports, and resources are perceived as most valuable by parents when preparing for their adolescents’ transition to adult care?
5. Which chronic illnesses or medical specialties have been most frequently studied in relation to parental support within the context of HCTs?
6. In what ways, if any, might cultural and demographic influences (e.g., race, ethnicity, socioeconomic status, immigration status, and geographical location) impact parental preparedness?
7. What recommendations have been made to enhance parental support and preparedness during transition?

### Inclusion criteria

The provisional inclusion and exclusion criteria for this study is informed by the population, concepts and context (PCC) framework as summarized in Table 1.

**Table 1.**
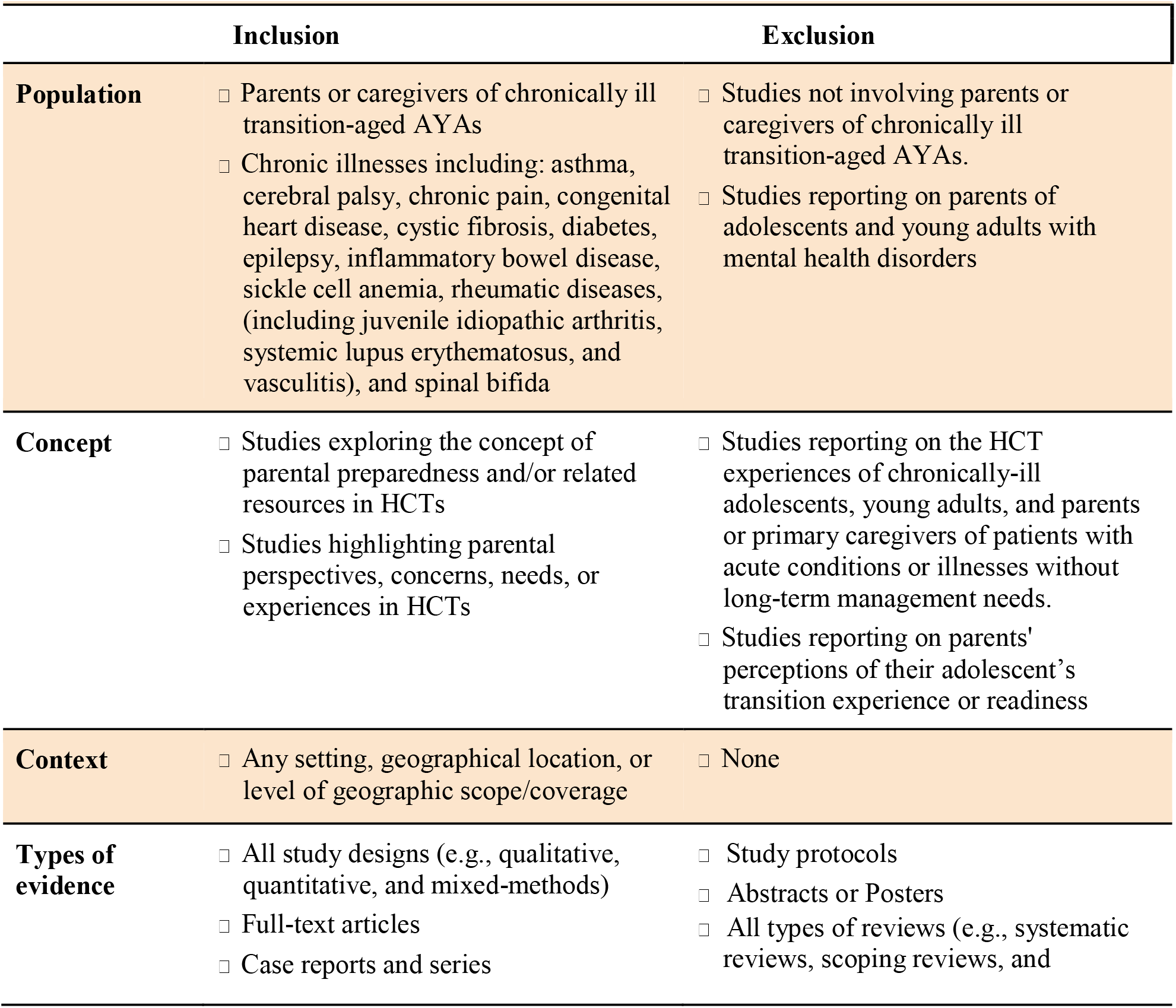
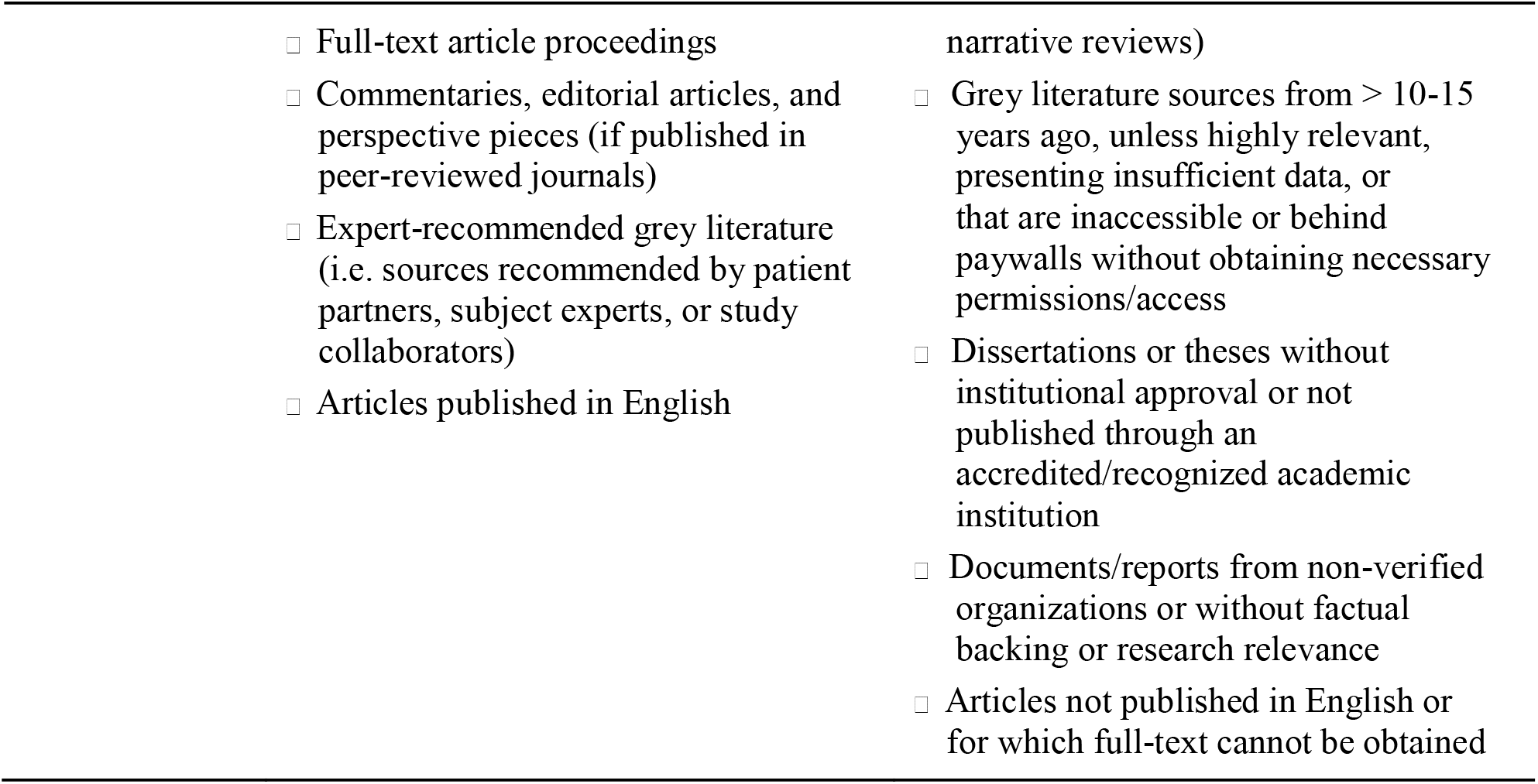
Review eligibility criteria based on study population, concept, context and types of evidence.

|  |  |  |
| --- | --- | --- |
|  | **Inclusion** | **Exclusion** |
| **Population** | □ Parents or caregivers of chronically ill transition-aged AYAs<br>□ Chronic illnesses including: asthma, cerebral palsy, chronic pain, congenital heart disease, cystic fibrosis, diabetes, epilepsy, inflammatory bowel disease, sickle cell anemia, rheumatic diseases, (including juvenile idiopathic arthritis, systemic lupus erythematosus, and vasculitis), and spinal bifida | □ Studies not involving parents or caregivers of chronically ill transition-aged AYAs.<br>□ Studies reporting on parents of adolescents and young adults with mental health disorders |
| **Concept** | □ Studies exploring the concept of parental preparedness and/or related resources in HCTs<br>□ Studies highlighting parental perspectives, concerns, needs, or experiences in HCTs | □ Studies reporting on the HCT experiences of chronically-ill adolescents, young adults, and parents or primary caregivers of patients with acute conditions or illnesses without long-term management needs.<br>□ Studies reporting on parents' perceptions of their adolescent's transition experience or readiness |
| **Context** | □ Any setting, geographical location, or level of geographic scope/coverage | □ None |
| **Types of evidence** | □ All study designs (e.g., qualitative, quantitative, and mixed-methods)<br>□ Full-text articles<br>□ Case reports and series | □ Study protocols<br>□ Abstracts or Posters<br>□ All types of reviews (e.g., systematic reviews, scoping reviews, and |

|  |  |
| --- | --- |
| □ Full-text article proceedings<br>□ Commentaries, editorial articles, and perspective pieces (if published in peer-reviewed journals)<br>□ Expert-recommended grey literature (i.e. sources recommended by patient partners, subject experts, or study collaborators)<br>□ Articles published in English | narrative reviews)<br>□ Grey literature sources from > 10-15 years ago, unless highly relevant, presenting insufficient data, or that are inaccessible or behind paywalls without obtaining necessary permissions/access<br>□ Dissertations or theses without institutional approval or not published through an accredited/recognized academic institution<br>□ Documents/reports from non-verified organizations or without factual backing or research relevance<br>□ Articles not published in English or for which full-text cannot be obtained |

#### Population

This review will consider all sources of evidence that report on parents or caregivers directly involved in the care of their chronically ill adolescents and their transition from pediatric to adult care. No limitations with regards to the parents’ or caregivers’ age, sex, marital status, or other demographic characteristics will be applied.

#### Concept

The primary concept examined in this scoping review is parental preparedness, describing the extent to which parents feel well equipped to support and navigate this shared transition from pediatric to adult care with their chronically ill AYAs, including parents’ perceptions of the successes and challenges influencing this process. Additionally, for the purposes of this review, we will follow CDC’s definition of chronic illnesses as “conditions that last at least one year, require ongoing medical care, and limit daily activities."<u>^28^</u>

#### Context

This scoping review is not limited to any particular context, including healthcare settings (e.g., hospitals, specialty clinics), communities (socioeconomic, ethnic, cultural communities), geographical setting (urban vs rural settings) or level of geographical scope/coverage (e.g., local, provincial, and national settings); thus considering all published literature on HCTs.

#### Types of evidence sources

This review will consider the inclusion of all transition-focused research employing any methodological approach, including qualitative, quantitative, or mixed-methods study design. Qualitative study designs may include phenomenology, grounded theory, ethnography, and narrative-based approaches in research. Quantitative studies may include observational (e.g., cohort, case-control, cross-sectional, descriptive studies), experimental (e.g., randomized controlled trials), and quasi-experimental (e.g. non-randomized controlled trials) study designs. All types of reviews (e.g., systematic reviews, scoping reviews, narrative reviews) and study protocols will be excluded, unless determined to be majorly pertinent to the subject topic, in which case their reference lists will be hand-searched to identify relevant sources. In addition, case series and individual case reports, while rare in studies exploring parental preparedness in HCTs, will be included. Inclusion of text and opinion papers (e.g., commentaries, editorials, and perspective pieces) will only be considered if published in an academic peer-reviewed journal. Grey literature sources recommended by patient partners, clinical and qualitative research experts, among other study collaborators will also be searched, including, but not limited to sources such as dissertations/doctoral theses, conference/event proceedings, and white papers (e.g., organizational reports, industry publications). Considering the exploratory nature of this scoping review, the inclusion of both scholarly and grey literature sources in the literature search will help ensure a comprehensive coverage of relevant findings, minimizing publication bias, enhancing rigor and transparency, and ultimately contribute to a more nuanced understanding of parental perceptions and experiences in HCTs.

### Stage 2: Identifying relevant studies

To ensure the development of a comprehensive search strategy, an iterative approach was employed in collaboration with an experienced librarian consultant at McMaster University (SC). A draft trial search was first performed in Ovid MEDLINE to identify relevant articles, key words, and index terms, based on the review’s primary objectives and research questions. The results of the preliminary search were reviewed to determine their overall relevance and comprehensiveness, following which subject-specific terminology relating to key terms such as, “parents", “adolescents and young adults", “healthcare transitions", and “chronic illness", identified from the titles and abstracts of pertinent literature were used to make informed revisions to the search strategy. The complete Ovid MEDLINE search strategy can be found in online supplemental appendix 3. The targeted search strategy will be adapted and applied in full across each of the five remaining databases, including CINAHL via EBSCOhost, APA PsycInfo via Ovid, Embase via Ovid, Web of Science Core Collection, and Sociological Abstracts via ProQuest, based on their respective indexing standards and naming conventions. Grey literature sources recommended by patient partners, subject experts, and study collaborators will also be searched and considered for inclusion in this review. Parent and patient partners involved in the study as both co-authors and lived experience experts offered valuable insights in incorporating controlled vocabulary by identifying key terms and phrases reflecting both their real-world understanding of healthcare transitions, as well as appropriate clinical and research terminology specific to this process. To ensure an exhaustive and robust review process, the reference list of all included studies will also be hand-searched to maximize the retrieval of relevant sources. Only reports published in English will be included. The search will include internationally published literature, but will be restricted to articles published in English. All literature searches will be thoroughly reviewed and finalized by an expert librarian consultant and information specialist.

### Stage 3: Study selection

All identified citations will be collated and uploaded into Covidence Systematic Review Software *(Veritas Health Innovation, Melbourne, Australia)*, and duplicates removed.<u>^29^</u> A two-stage study selection process will be implemented, involving the participation of three to four reviewers. In stage 1, two reviewers (PM, CT) will first perform title and abstract review of all results to ensure consistent application of the pre-defined inclusion and exclusion criteria, following which any conflicts will be resolved by a third and/or fourth reviewer (SM, KF). In stage 2, two reviewers (PM, CT) will independently perform a comprehensive full-text review of the remaining articles. Only articles determined to fulfill the inclusion criteria by both reviewers will be included in the review. Any discrepancies or a lack of consensus in the study selection or review process will be resolved by a third and/or fourth reviewer (SM, KF).

### Stage 4: Data Extraction

Data charting will be performed utilizing a standardized data extraction tool adapted from the data charting template developed by the JBI manual for evidence synthesis.^25^ To help ensure high inter-reviewer reliability and charting consistency the draft charting form will first be piloted by two reviewers independently on a random sample of included studies at the review stage. If poor agreement is found, the data extraction tool will be revised iteratively, and the pilot exercise subsequently repeated. In following the PCC framework, the data charted in the review will include specific details about population, concept, context, study design/methodology, and parental perceptions and experiences in HCTs. Refer to Table 2 for more details on the data charting of review-specific fields and variables. If needed, the corresponding authors of included papers will also be contacted to inquire about or request any missing or additional data. Recognizing that the principal focus of our scoping review is to map any available evidence pertinent to parental perceptions and experiences in HCTs, a formal critical appraisal of included literature will not be performed.

**Table 2.**
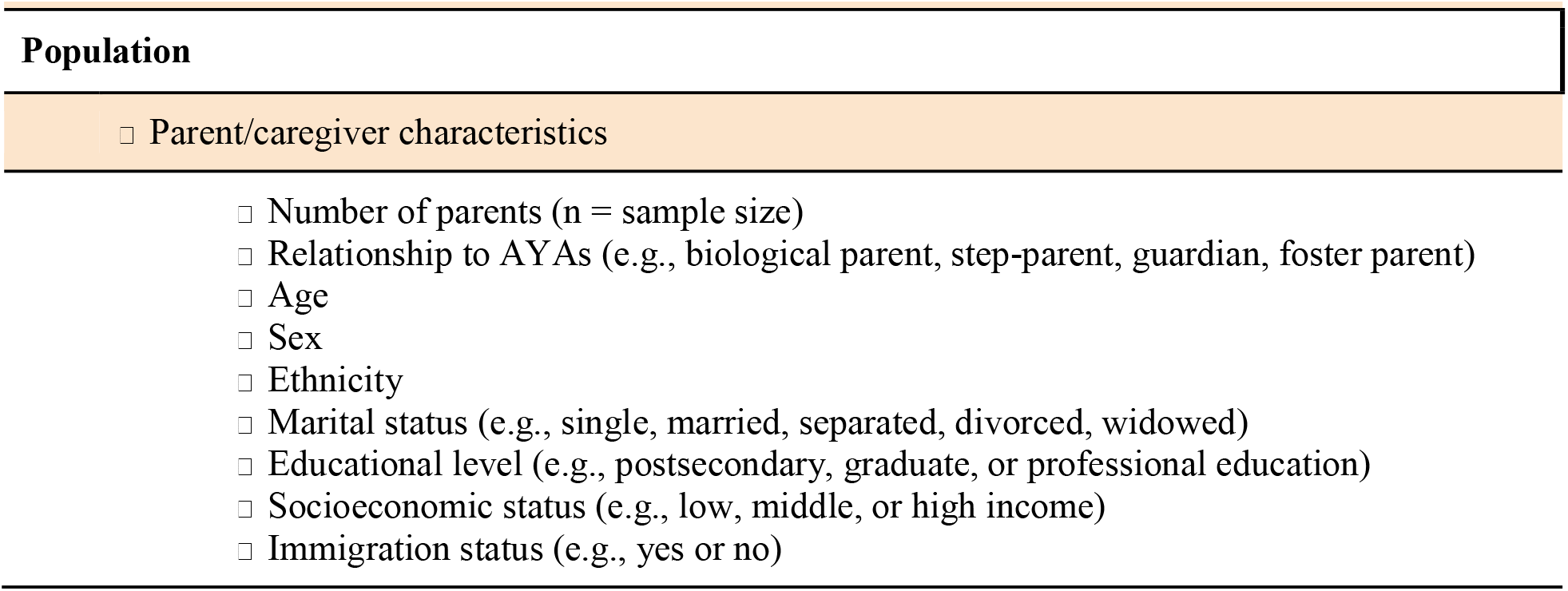
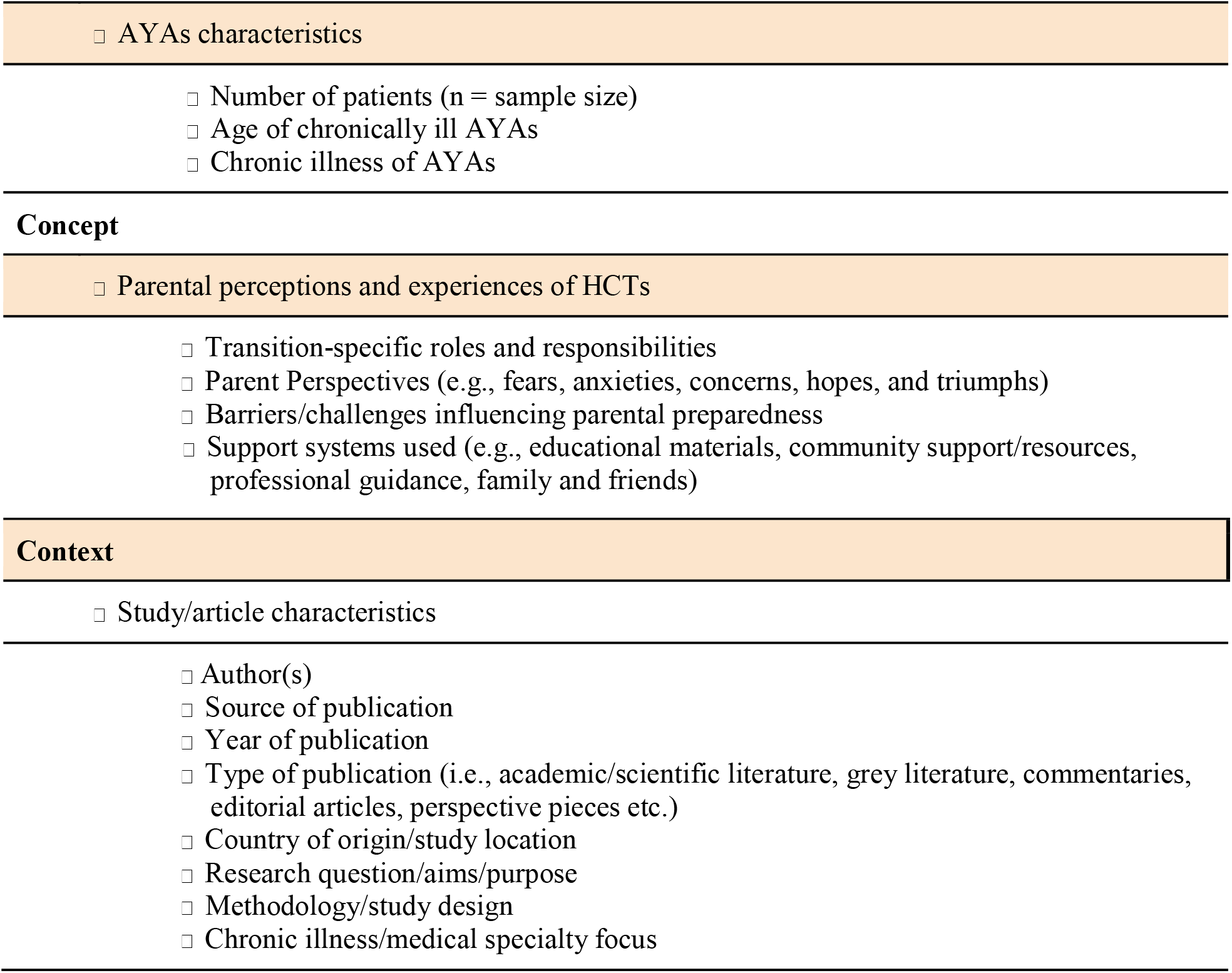
Data charting fields for the scoping review.

| Table 2. Data charting fields for the scoping review. |
| --- |
| Population |
| □ Parent/caregiver characteristics |
| □ Number of parents (n = sample size)<br>□ Relationship to AYAs (e.g., biological parent, step-parent, guardian, foster parent)<br>□ Age<br>□ Sex<br>□ Ethnicity<br>□ Marital status (e.g., single, married, separated, divorced, widowed)<br>□ Educational level (e.g., postsecondary, graduate, or professional education)<br>□ Socioeconomic status (e.g., low, middle, or high income)<br>□ Immigration status (e.g., yes or no) |
| □ AYAs characteristics |
| □ Number of patients (n = sample size)<br>□ Age of chronically ill AYAs<br>□ Chronic illness of AYAs |
| **Concept** |
| □ Parental perceptions and experiences of HCTs |
| □ Transition-specific roles and responsibilities<br>□ Parent Perspectives (e.g., fears, anxieties, concerns, hopes, and triumphs)<br>□ Barriers/challenges influencing parental preparedness<br>□ Support systems used (e.g., educational materials, community support/resources, professional guidance, family and friends) |
| **Context** |
| □ Study/article characteristics |
| □ Author(s)<br>□ Source of publication<br>□ Year of publication<br>□ Type of publication (i.e., academic/scientific literature, grey literature, commentaries, editorial articles, perspective pieces etc.)<br>□ Country of origin/study location<br>□ Research question/aims/purpose<br>□ Methodology/study design<br>□ Chronic illness/medical specialty focus |

### Stage 5: Collating, review and reporting of results

To ensure transparency and reproducibility in the review process, study selection and screening of all sources will be reported using a PRISMA-ScR flow diagram.^26^ Charted data will be analyzed qualitatively (e.g., thematic or framework analysis) and quantitatively (e.g., descriptive statistics, frequency analysis, data visualization) using the Statistical Package for Social Sciences (SPSS) version 28, where applicable.

### Stage 6: Consultation

As co-leads, patient partners were actively involved in developing the search strategy as they shared their insights on which key terms were most representative of their lived experiences. Additionally, in tandem with level 1 screening, patient partners and study collaborators will be asked to share any grey literature sources they consider to be pertinent to our review’s primary objectives and research questions. Short informal semi-structured interviews will also be organized with research experts recruited through the team’s personal and professional networks to further validate preliminary findings and identify any gaps in the scope, analysis, and/or interpretation of findings.

### Study Status

At the time of this protocol’s submission, the literature searches across all databases have been completed and await screening.

## Data Availability

All data generated for this scoping review protocol are publicly available online and have been appropriately cited within the manuscript.

## Supplementary Information

### Supplemental Appendix 1

PRISMA-ScR checklist

### Supplemental Appendix 2

GRIPP2 checklist

### Supplemental Appendix 3

Complete Ovid MEDLINE search strategy

