## Supplemental Appendix 2 for "Addressing the Needs and Identifying Supports for Parents of Chronically Ill Adolescents and Young Adults in their Shared Transition from Pediatric to Adult Care: A Scoping Review Protocol"

GRIPP2 short form

| **Section and topic** | **Item** | **Reported on page No** |
| --- | --- | --- |
| 1: Aim | Report the aim of PPI in the study | 5 |
| 2: Methods | Provide a clear description of the methods used for PPI in the study | 5 |
| 3: Study results | Outcomes—Report the results of PPI in the study, including both positive and negative outcomes | N/A |
| 4: Discussion and conclusions | Outcomes—Comment on the extent to which PPI influenced the study overall. Describe positive and negative effects | N/A |
| 5: Reflections/critical perspective | Comment critically on the study, reflecting on the things that went well and those that did not, so others can learn from this experience | N/A |

Staniszewska S, Brett J, Simera I, et al. GRIPP2 reporting checklists: tools to improve reporting of patient and public involvement in research. *BMJ*. 2017;358:j3453. Published 2017 Aug 2. doi:10.1136/bmj.j3453
