## Supplemental Appendix 3 for "Addressing the Needs and Identifying Supports for Parents of Chronically Ill Adolescents and Young Adults in their Shared Transition from Pediatric to Adult Care: A Scoping Review Protocol"

**Database: Ovid MEDLINE(R) ALL <1946 to December 18, 2024>**
**Search Strategy:**
**1**  parents/ or fathers/ or mothers/ or single parent/ or caregivers/ or legal guardians/ or family support/ or maternal behavior/ or parenting/ or paternal behavior/ or parent-child relations/ or father-child relations/ or mother-child relations/ (258638)
**2**  (parent* or father? or mother? or caregiv* or guardian? or family or families).tw,kf,kw. (1926657)
**3**  1 or 2 (1975555)
**4**  adolescent/ or young adult/ or minors/ or Adolescent Health/ or adolescent health services/ or adolescent medicine/ or Adolescent Nutritional Physiological Phenomena/ or Pediatrics/ (2850252)
**5**  (adolescen* or teen? or teenage* or transition-aged or juvenile? or young person? or young people or young adult? or puberty or pubescen* or youth? or pre-adult* or young adult* or emerging adult* or p?ediatric*).tw,kf,kw. (1192160)
**6**  4 or 5 (3439148)
**7**  exp Asthma/ (147494)
**8**  (asthma or lung allerg*).tw,kf,kw. (181378)
**9**  Cerebral Palsy/ (24867)
**10**  (((cerebral or brain or central) adj (palsy or paralysis)) or encephalopathia infantilis or (spastic? adj1 diplegia)).tw,kf,kw. (30246)
**11**  Chronic Pain/ (25982)
**12**  (chronic adj2 pain).tw,kf,kw. (79703)
**13**  exp Heart Defects, Congenital/ (175160)
**14**  (congenital adj (heart or cardiac) adj (disease or distress)).tw,kf,kw. (38654)
**15**  Cystic Fibrosis/ (41479)
**16**  ((cystic adj2 (fibrosis or disease)) or fibrocystic disease or mucoviscidosis).tw,kf,kw. (60748)
**17**  diabetes mellitus/ or diabetes mellitus, type 1/ (233057)
**18**  (diabetes or diabetic).tw,kf,kw. (841183)
**19**  exp Epilepsy/ (131602)
**20**  epilep*.tw,kf,kw. (177008)
**21**  exp Inflammatory Bowel Diseases/ (103942)
**22**  (((ulcer* or mucosal) adj2 (colitis or colorectitis or proctocolitis)) or (Crohn* adj (disease or morbus))).tw,kf,kw. (93011)
**23**  inflammatory bowel disease?.tw,kf,kw. (72717)
**24**  Anemia, Sickle Cell/ (25250)
**25**  ((sickle cell adj (an?emia or disease)) or drepanocytemia or drepanocytosis or drepanocytic an?emia or hbs disease or h?emoglobin s disease).tw,kf,kw. (26771)
**26**  exp rheumatic diseases/ (272908)
**27**  ((rheumat* adj2 (disease or syndrome or arthrit* or polyarthrit* or chronic)) or ((chronic or juvenile) adj2 (arthrit* or polyarthrit*))).tw,kf,kw. (165163)
**28**  exp Lupus Erythematosus, Systemic/ (70045)
**29**  lupus.tw,kf,kw. (96285)
**30**  exp Vasculitis/ (107025)
**31**  ((vascul* adj2 (inflammat* or lesion* or syndrome* or disease or disorder*)) or ang?itis).tw,kf,kw. (88321)
**32**  exp Spinal Dysraphism/ (9201)
**33**  (spinal dysraphi* or spina bifida or cleft spine? or open spine? or rachischis*).tw,kf,kw. (10511)
**34**  or/7-33 (2368157)
**35**  transitional care/ or transition to adult care/ or "continuity of patient care"/ or patient transfer/ or patient handoff/ (35225)
**36**  ((leav* or exit*) adj2 p?ediatric care).tw,kf,kw. (19)
**37**  "p?ediatric to adult".tw,kf,kw. (11217)
**38**  ((enter* or entry* or move? or moving) adj4 adult care).tw,kf,kw. (55)
**39**  ((care or healthcare) adj3 (transition or transfer or coordinat* or continuum or continuity)).tw,kf,kw. (43626)
**40**  or/35-39 (78365)
**41**  3 and 6 and 34 and 40 (1123)
